# Single-Particle Counting Based on Digital Plasmonic Nanobubble Detection for Rapid and Ultrasensitive Diagnostics

**DOI:** 10.1101/2021.02.18.21252027

**Authors:** Yaning Liu, Haihang Ye, HoangDinh Huynh, Peiyuan Kang, Chen Xie, Jeffrey S. Kahn, Zhenpeng Qin

**Author notes:** Co-first authors (Yaning Liu, Haihang Ye).

## Abstract

Rapid and sensitive diagnostics of infectious diseases is an urgent and unmet need as evidenced by the COVID-19 pandemic. Here we report a novel strategy, based on DIgitAl plasMONic nanobubble Detection (DIAMOND), to address these gaps. Plasmonic nanobubbles are transient vapor bubbles generated by laser heating of plasmonic nanoparticles and allow single-particle detection. Using gold nanoparticles labels and an optofluidic setup, we demonstrate that DIAMOND achieves a compartment-free digital counting and works on homogeneous assays without separation and amplification steps. When applied to the respiratory syncytial virus diagnostics, DIAMOND is 150 times more sensitive than commercial lateral flow assays and completes measurements within 2 minutes. Our method opens new possibilities to develop single-particle digital detection methods and facilitate rapid and ultrasensitive diagnostics.

**One Sentence Summary:** Single-particle digital plasmonic nanobubble detection allows rapid and ultrasensitive detection of viruses in a one-step homogeneous assay.

## Main Text

The ability to rapidly detect diseases with high precision is of paramount importance as evidenced by the current COVID-19 pandemic *(1, 2)*. Digital assays have been a remarkable conceptual advance over the past two decades due to their single-molecule detection and absolute quantification *(3, 4)*. They partition the analytes into microwells or emulsion droplets as small compartments for independent signal amplification and digital counting, leading to the sensitivity enhancement by up to 10^3^-fold over the conventional assays (i.e., enzyme-linked immunosorbent assay and polymerase chain reaction) *(3, 4)*. Despite those advantages, digital assays have suffered from complex assay operations. Such paradigms prompt further innovations that develop various digital sensing platforms based on micro/nano-particles *(5-7)*. With its capability of examining individual particles’ changes upon recognizing target molecules, single-particle detection holds great potential to simplify digital assays *(8-10)*. Examples of single-particle digital assays include bright/dark-field imaging *(11)*, interferometric *(12)* or fluorescent imaging *(13, 14)*, surface-enhanced Raman scattering *(15)*, surface plasmon resonance microscopy imaging *(16)*, and particle mobility tracking *(17-19)*. However, current techniques rely on cumbersome particle purification and advanced imaging that inevitably limit their widespread use.

Herein, we report a novel strategy for simplified single-particle digital assay based on DIgitAl plasMONic nanobubble Detection (DIAMOND, see **Table S1** for comparison with other assays). Plasmonic nanobubbles (PNBs) refer to the vapor bubbles generated by short laser pulse excitation of plasmonic nanoparticles (NPs) and amplify their intrinsic scattering for the detection by a secondary probe laser *(20-24)*. They have lifetime that lasts nanoseconds and are sensitive to the physical parameters of NPs such as sizes, shape, concentration, and clustering state *(20-22)*. Taking advantage of these unique properties, we designed an optofluidic setup to flow the NP suspension in a micro-capillary (**Fig. 1A** and **Fig. S1**). The focused laser beam probes a microscale “virtual compartment” of about 16 pL and detects the PNB generation from single particles. Since PNBs are transient events, there is no cross-talk between laser pulses, which allows “on” and “off” signal counting in a compartment-free manner (**Fig. 1B**). We demonstrated robust single NP counting and sensitive detection of large particles in a strong background of small particles (1 in 240). We then implemented DIAMOND in a homogeneous assay that uses plasmonic gold NPs (AuNPs) as labels without additional separation or amplification steps. Using silica (SiO_2_) beads as targets, we demonstrated that DIAMOND has sub-femtomolar detection limit and provides absolute quantification. When applied to detect respiratory syncytial virus (RSV), DIAMOND provided a 150-fold sensitivity enhancement over the state-of-the-art lateral flow assay (LFA) and sample-to-answer time within 2 minutes. Therefore, DIAMOND opens new possibilities to develop separation-, amplification-, and compartment-free single-particle digital assays and facilitate rapid and ultrasensitive diagnostic platforms.

**Fig. 1.**
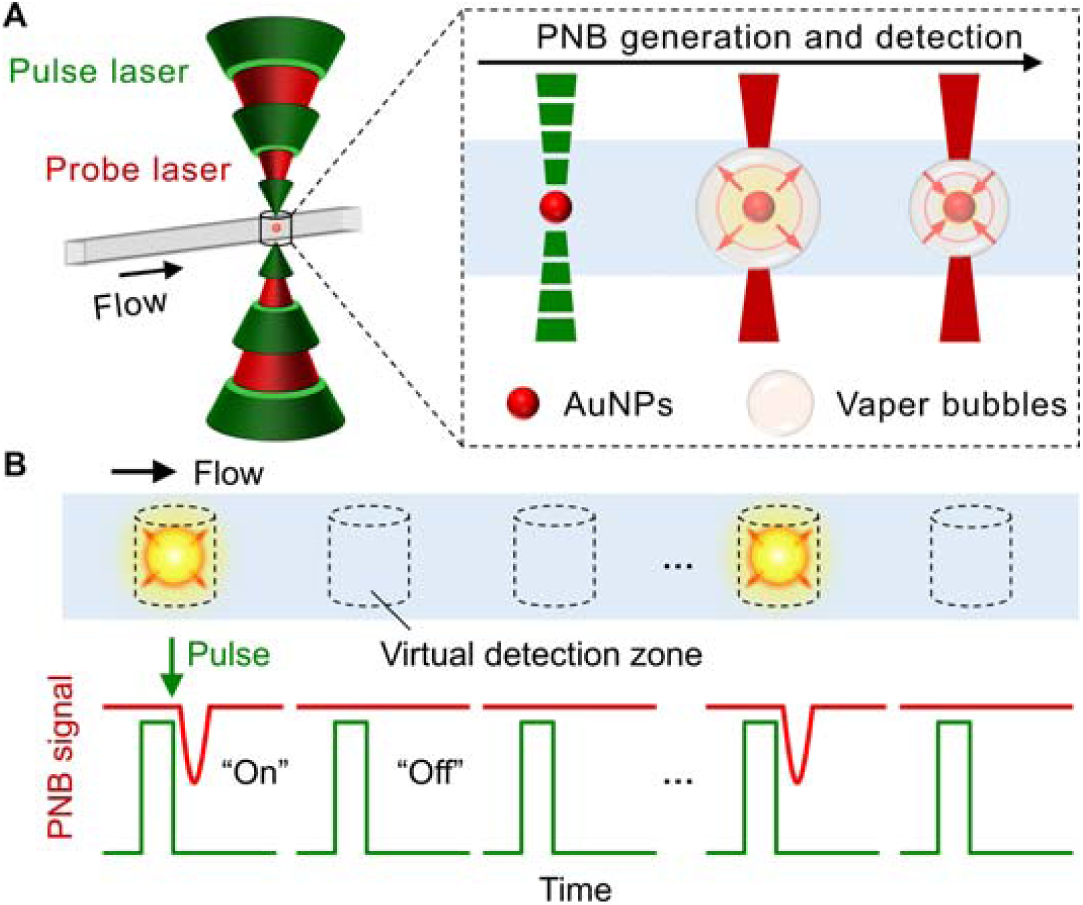
The schematics illustration for the concept of DIAMOND. (A) The spectroscopy-based signal generation and detection. The gold nanoparticles (AuNPs) as labels are used for the generation of the plasmonic nanobubbles (PNBs) by short laser pulses and subsequently detected by a secondary probe laser due to the optical scattering. (B) The detection principle based on optofluidic scanning of a sample flowing through. The “on” and “off” refer to the positive and negative PNB signals representing for the presence or absence of targets.

## Results

We first evaluated the ability of DIAMOND for single NP detection. Serial aqueous dilutions of 75 nm AuNP (as characterized in **Fig. S2, 3** and **Table S2**) were prepared for the DIAMOND tests. For simplicity, we converted the particle concentration into the expected average number (λ) of AuNPs per detection zone. **Fig. 2A** and **Fig. S4** show the representative testing results and the corresponding Poisson distributions for the given λ, respectively. Due to the variation of actual AuNP number (*k*) per detection zone, it led to discrete PNB signals with fluctuated intensity. In the case of λ=0.04, the positive PNB signals could be satisfactorily discriminated from the baseline (**Fig. 2A**). According to the Poisson statistics, each of signal drop refers to a single AuNP being detected (**Fig. S4D**), confirming the capability of DIAMOND in single-NP detection. To correlate the PNB signals with AuNP concentration, we calculated the frequencies (*f*) of positive PNB signals in each trace as “on” signals and plotted the *f*_on_ as a function of λ (**Fig. 2B**). A high correlation coefficient (*R*^2^=0.998) indicates the AuNP quantification by counting the frequency of “on” signal. Furthermore, we found a linear relationship (slope=1.012, *R*^2^=0.999, **Fig. 2C**) between the theoretical probability (*P*) predicted by Poisson statistics and the experimental *f*_on_, suggesting that DIAMOND is a calibration-free technique and allows absolute quantification.

**Fig. 2.**
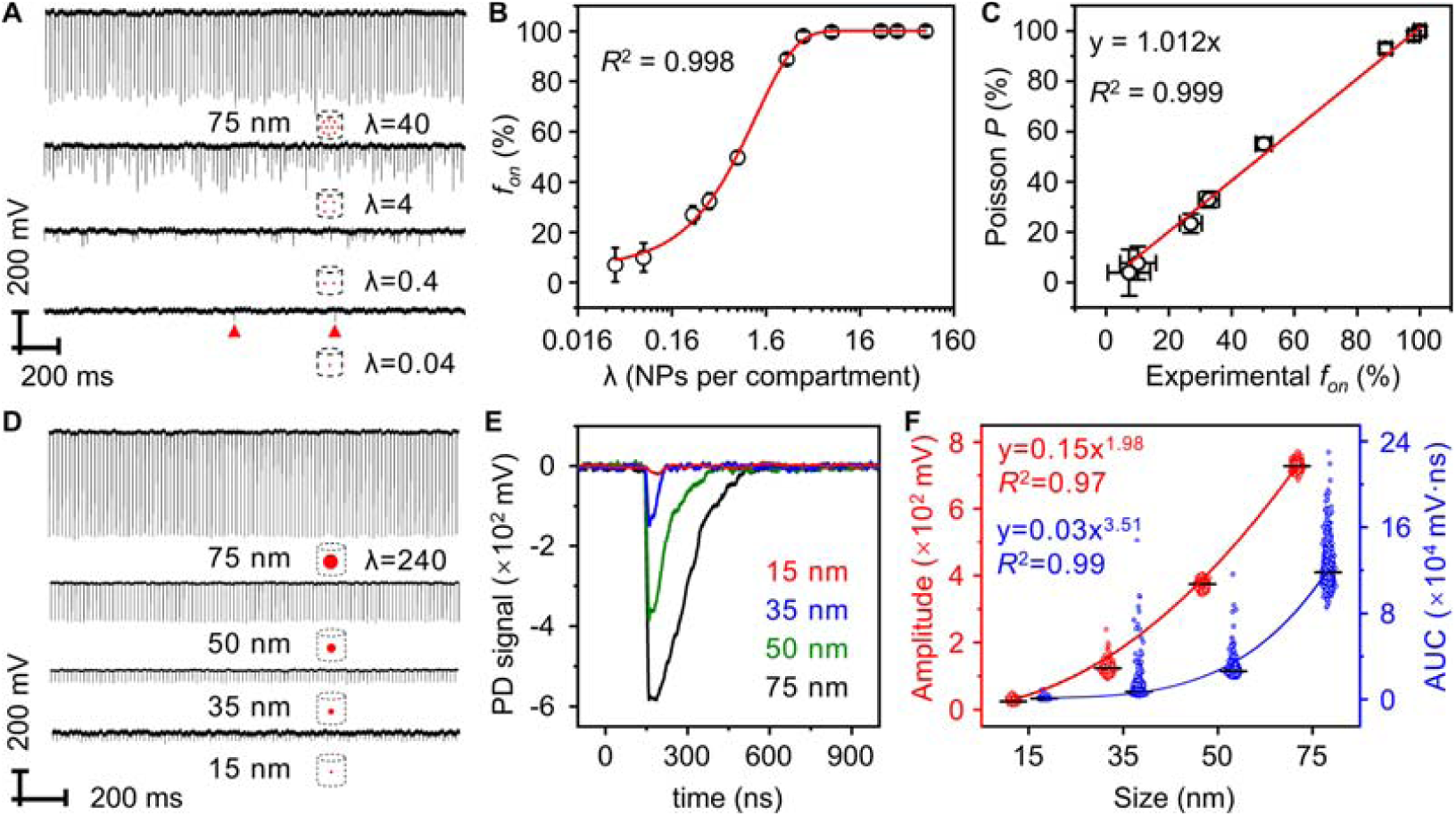
Detection of single AuNPs and differentiation of NP size by DIAMOND. (A) Representative PNB signal traces (100 pulses) for 75 nm AuNP suspensions with different particle concentrations. Schematics represent the decreased number (λ) of AuNPs per detection. Red triangles mark the positive PNB signals. (B) Quantification of AuNP concentrations by plotting the frequency count (*f*_on_) against λ. (C) Correlation between experimental *f*_on_ and the theoretical probability (*P*) based on Poisson statistics. (D) Representative PNB signals traces (100 pulses) for AuNP suspensions with different sizes. Schematics represent the increasing AuNP sizes and same AuNP number per detection. (E) Single PNB signals extracted from (D). PD is photodetector. (F) Correlations between the amplitude and AUC of PNB signals as a function of AuNP size. Black lines denote the statistical average values.

Next, we used DIAMOND to detect AuNPs of different sizes under the same concentration. AuNP suspensions of 15 nm, 35 nm, 50 nm, and 75 nm (**Fig. S2, 3** and **Table S2**) were prepared for the test. **Fig. 2D** shows the representative PNB signal traces. Due to the large number of AuNPs per detection zone (λ=240), consistent PNB signals were observed in each laser pulse. **Fig. 2E** shows representative PNB signals taken from each case and suggests that larger NP size leads to larger PNB signals. To quantify this relationship, we extracted the values of amplitude and area-under-the-curve (AUC) from each PNB signal (**Fig. S5**) and plotted their distribution profiles as a function of AuNP sizes (**Fig. 2F**). They could be fitted with 1.98- and 3.51-order dependences to the NPs’ size, respectively. This seems to agree with the fact that the PNB generation is related to the amount of heat generation and absorption cross-section of AuNPs *(25, 26)*. The good fitting indicates the accurate size determination by measuring the amplitude and AUC of PNB signals.

We then evaluated the ability of DIAMOND for heterogeneity identification in a population of NPs. 15 nm and 75 nm AuNP suspensions and their mixture were tested. **Fig. 3A** shows the representative PNB signal traces taken from those samples. The large intensity variation of PNB signals observed in the mixture sample is due to the addition of 75 nm AuNPs. To sort those specific signals resulting from 75 nm AuNPs as heterogeneities, we developed a three-step analysis protocol. Firstly, we extracted the amplitude and AUC values of each PNB signal as indexes for quantification. **Fig. 3B** shows the corresponding results in bivariate, where each scatter represents a single PNB signal. Secondly, we fitted normal distributions to the amplitude and AUC of 15 nm AuNPs as a reference and calculated the threshold (*T*) as 5 standard deviations (σ) above the mean (μ). As such, the two thresholds covered over 99.9% of the scatters (**Fig. 3B**, case *i*). Lastly, assigning the thresholds to the mixture sample allows data sorting, where the scatters above the thresholds refer to the positive or “on” PNB signals resulting from co-existence of 15 nm and 75 nm AuNPs and *vice versa* (**Fig. 3B**, case *ii*). For a direct comparison, we calculated the frequencies of “on” signals (*f*_on_) from 75 nm AuNPs alone (**Fig. 3B**, case *iii*) and benchmarked them with the Poisson probability. The agreement between experimental result and theoretical prediction (**Fig. 3C**) indicates that our method provides accurate and absolute quantification of large NPs from a strong background of small ones (i.e., 1 in 240). Clearly, DIAMOND is able to detect heterogeneity without additional separation step.

**Fig. 3.**
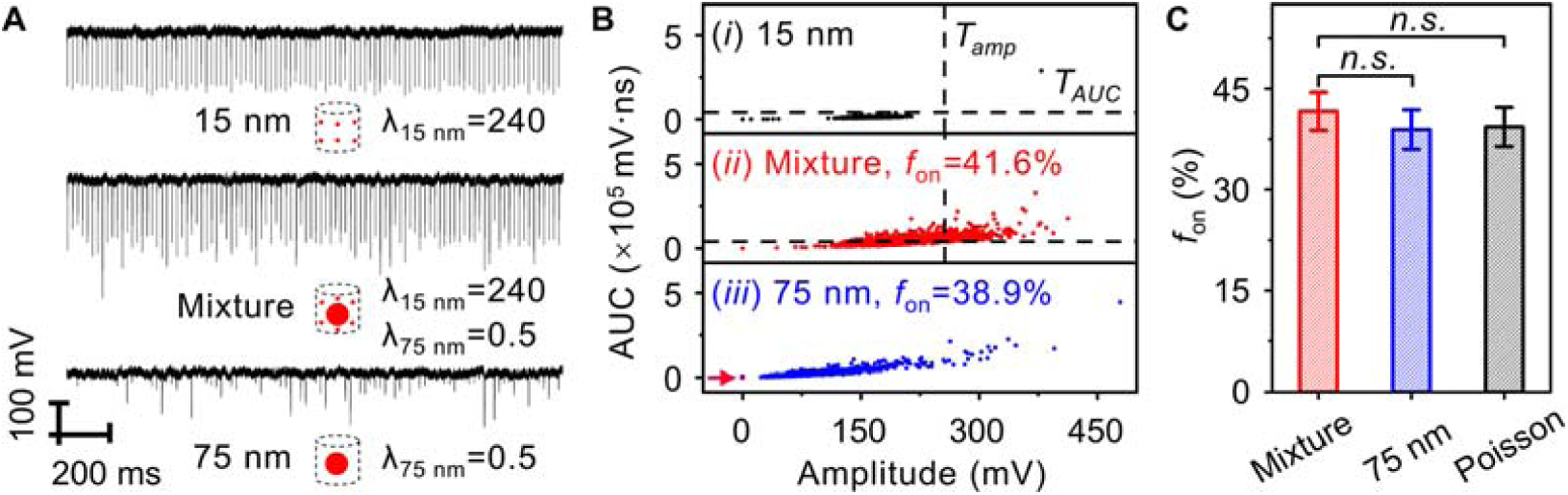
Identification of heterogeneity by DIAMOND. (A) Representative PNB signals traces (100 pulses) for 15 nm and 75 nm AuNPs and their mixture. Schematics represent the sample information. (B) Bivariate plots of amplitude and AUC extracted from 3,000 pulses for the three samples in (A). The dashed lines in cases (*i*) and (*ii*) indicate the same thresholds of amplitude (*T*_amp_) and AUC (*T*_AUC_). The red arrow in case (*iii*) highlights scatters at 0. (F) Bar plot of experimental frequencies (*f*_on_) as determined in (B) for cases (*ii*) and (*iii*) and theoretical probability predicted by Poisson statistics. n.s. stands for no significant difference (*p*-value>0.05). The error bars for experimental *f*_on_ indicate the standard deviations of three independent measurements and for theory, it is the Poisson noise-associated coefficient of variation 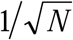, where *N* is the number of counted “on” signals.

We further evaluated the feasibility of implementing DIAMOND for the homogeneous assay. Homogeneous assays are simple and one-step sensing methods that require minimal sample handling and are therefore promising for rapid detection *(28)*. To validate our hypothesis in a model system, we used the AuNPs as probes to detect SiO_2_ beads (**Fig. 4A**). **Fig. 4B** shows the transmission electron microscopy (TEM) image of the as-prepared product, where a core-satellites structure was formed with AuNPs fully covering the surface of SiO_2_ beads. **Fig. 4C** shows representative PNB signal traces and **Fig. 4D** shows the bivariate plot taken from serial assay solutions. Similarly, the thresholds (*T*=*μ*+5□*σ*) of amplitude and AUC were calculated from the control group (highlighted in dashed lines) and assigned for “on” signal counting. The *f*_on_ were then plotted as a function of λ for SiO_2_ beads (**Fig. 4E**). A linear relationship (*R*^2^=0.99) in the range of 0.0016-0.16 was observed and the limit of detection (*LOD*) was calculated to be 0.0028, equivalent to 1.75×10^5^ mL^-1^ or 290 aM (inset of **Fig. 4E**). Such a detection limit is ∼570-fold lower than the colorimetric detection (**Fig. S6**). Furthermore, DIAMOND allows absolute quantification for homogeneous assays, as indicated by the linear correlation (slope=0.995, *R*^2^=0.99) between the background-subtracted frequency (*f*_on_’) and the Poisson probability (**Fig. 4F**).

**Fig. 4.**
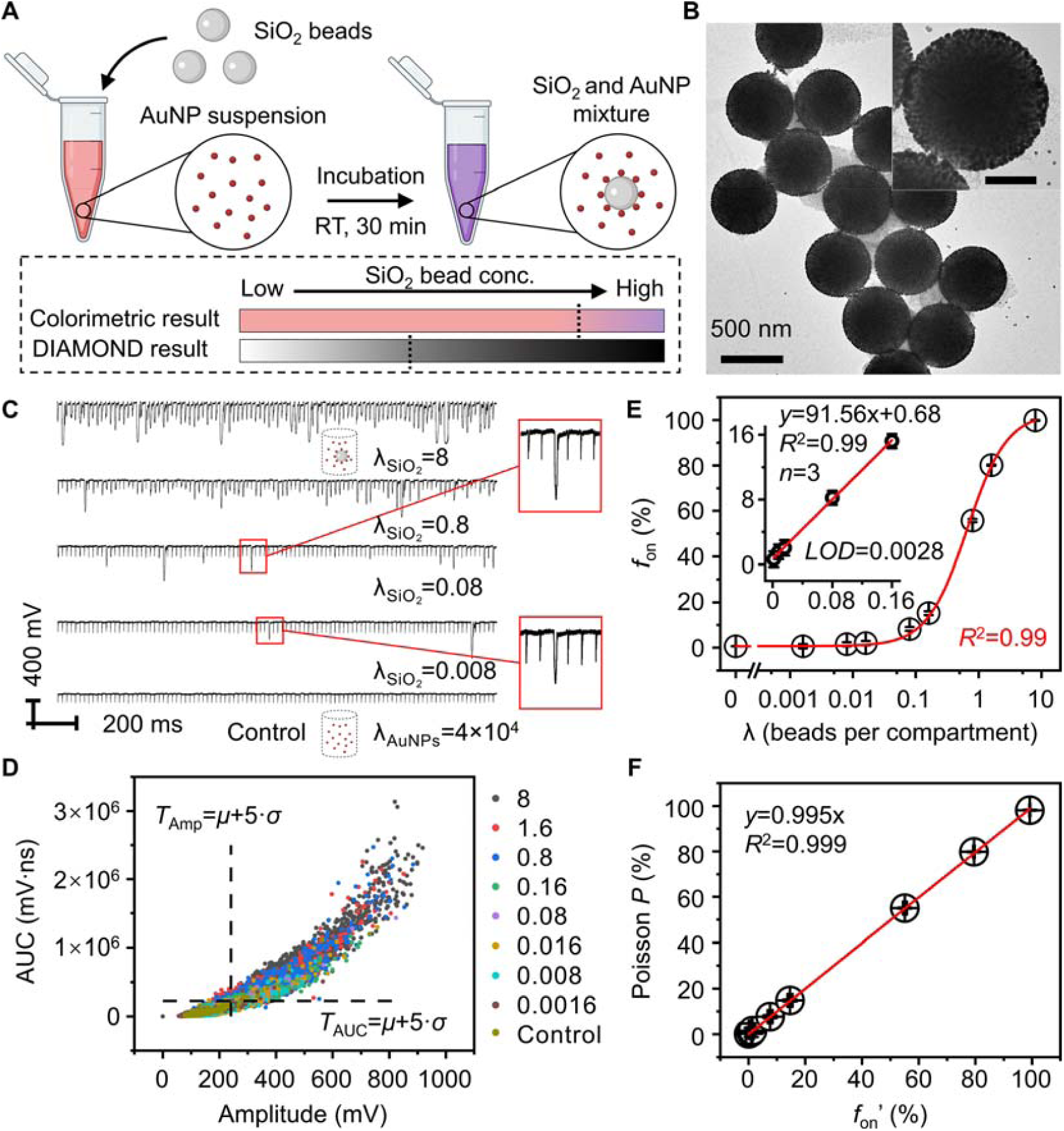
Detection of SiO_2_ beads in a homogeneous assay by DIAMOND. (A) Schematic of a homogeneous assay of SiO_2_ beads by AuNPs as a pair of targets and probes at room temperature (RT). Lower panel shows that when bead concentrations are insufficient to induce the color change, DIAMOND can detect the PNB signals. (B) TEM image of SiO_2_-AuNPs conjugates. Scale bar in inset is 200 nm. (C) Representative PNB signal traces (100 pulses) for the assay solutions. Schematics represent the assay information that show the different λ of SiO_2_ beads and the same λ of AuNPs. (D) Bivariate plot of amplitude and AUC extracted from 3,000 pulses for the assay solutions with different λ of SiO_2_ beads. Dashed lines indicate the positions of thresholds calculated from the control sample. (E) Quantification of SiO_2_ bead concentration as a function of frequency counting (*f*_on_). Error bars indicate the standard deviations of three independent measurements, and the *LOD* was calculated as 3 standard deviations of the control dividing the slope of regression line. (F) Correlation between the background-subtracted frequency (*f*_on_’) and Poisson probability (*P*).

Finally, we applied DIAMOND for the rapid detection of RSV. RSV is the major respiratory pathogen that accounts for up to 74,500 deaths in 2015 globally in children below age 5 *(28)*. We chose Synagis (Palivizumab) as the detection antibody and conjugated it on the AuNPs through 3,3’-dithiobis (sulfosuccinimidyl propionate, DTSSP) as a crosslinker (**Fig. S7**). The as-prepared probes could specifically target the fusion proteins on the RSV surface at the room temperature and were ready for detection (**Fig. 5A**). **Fig. 5B** shows TEM images of AuNP-based probes associated with RSV particles, indicating effective probe binding with the target. **Fig. S8** and **Fig. 5C** show the colorimetric detection of RSV (*LOD*: 3.6×10^4^ PFU/mL) and reveal the good detection specificity of AuNP probes towards RSV, among different respiratory viruses including Parainfluenza viruses (PIV), Influenza A (IVA), Human metapneumovirus (hMPV). For a direct comparison, we benchmarked the homogeneous immunoassay against the commercially available LFA kit for RSV, which has an *LOD* of 1.6×10^4^ PFU/mL (**Fig. S9**, BinaxNOW, Abbott). We then carried out DIAMOND for RSV detection. Following the same protocol as discussed above, we set the thresholds based on the control group (**Fig. 5D**) and assigned them for *f*_on_ counting (**Fig. 5E**). In particular, a linear relationship (*R*^2^=0.995) in the range of 10^2^-10^4^ PFU/mL was observed and the *LOD* was calculated to be 108 PFU/mL (inset of **Fig. 5E**). Evidently, DIAMOND achieved a sensitivity enhancement of ∼333-fold over the colorimetric result of homogeneous immunoassay and ∼150-fold over the commercial LFA.

**Fig. 5.**
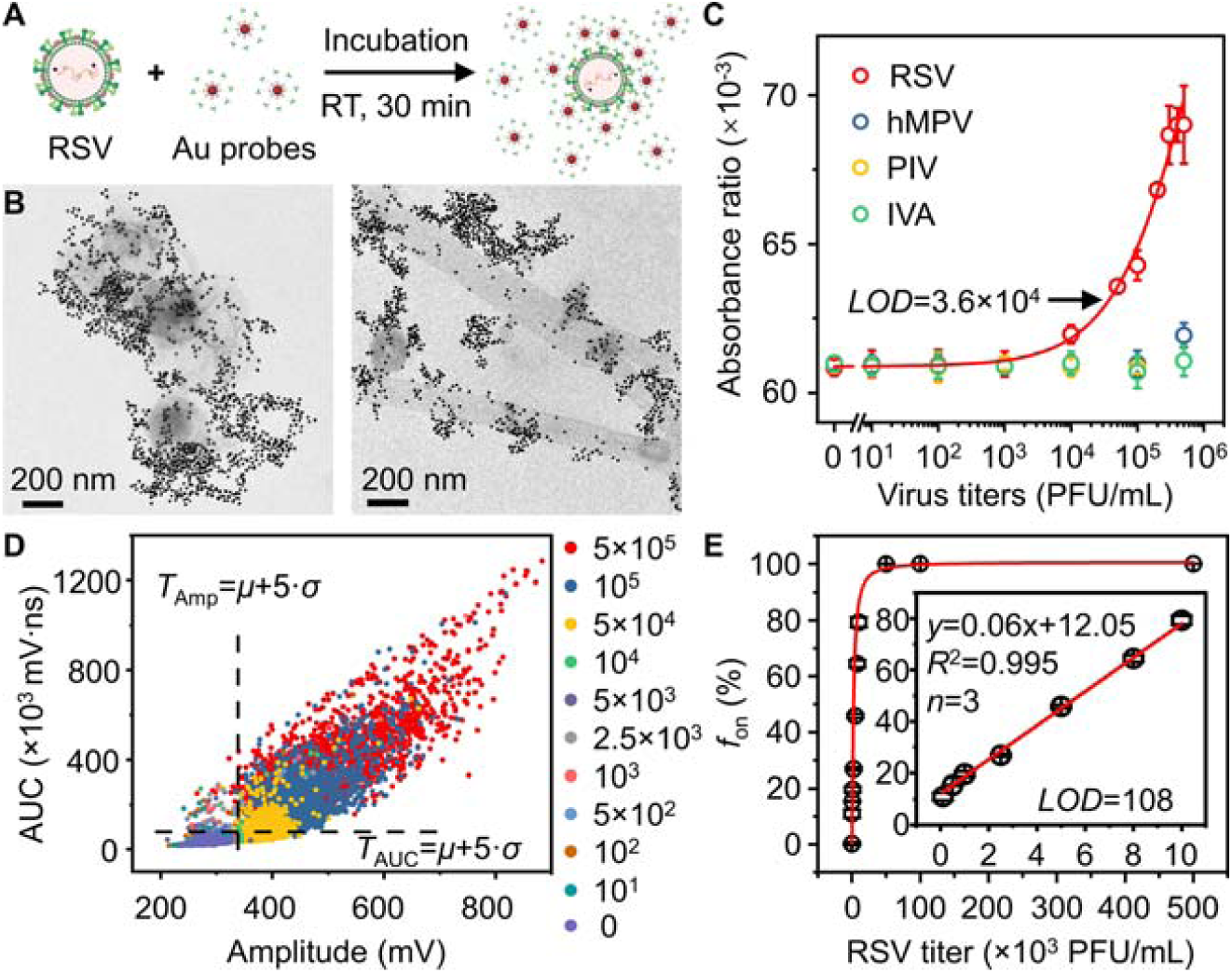
Detection of respiratory syncytial virus (RSV) in a one-step homogenous immunoassay by DIAMOND. (A) The schematic illustration of a homogeneous immunoassay for RSV utilizing antibody-conjugated AuNPs as probes at room temperature (RT). (B) TEM images of AuNP probes targeting RSV that shows different morphologies. (C) Colorimetric analysis of the AuNP-based homogeneous immunoassay of different respiratory viruses. hMPV is Human metapneumovirus, PIV is Parainfluenza viruses, and IVA is Influenza A. (D) Bivariate plot of amplitude and AUC extracted from 3,000 pulses for the assay solutions with different RSV titers (PFU/mL). Dashed lines indicate the positions of thresholds calculated from the control sample. (E) Quantification of RSV titers as a function of frequency counting (*f*_on_). Inset shows the linear detection range. Error bars in (C) and (E) indicate the standard deviations of three independent measurements, and the *LOD* was calculated as 3 standard deviations of the control divided by the slope of regression line.

To demonstrate the potential clinical use, we applied DIAMOND to quantify RSV in the transporting medium used to collect nasal swab samples. RSV at concentrations of 500-4,500 PFU/mL (as measured by end-point titration in viral culture) was spiked in the medium for testing. As summarized in **Table 1**, the recovery rate for DIAMOND in examining the five RSV-spiked samples was calculated to be 94.4-119%. The coefficient of variation (*CV*, n=6) increased from ∼6% to ∼20% with decreasing RSV concentration, as expected when approaching the *LOD*. These data demonstrated that the DIAMOND-coupled homogeneous immunoassay allows sensitive analysis of viral samples in complex matrices (i.e., transporting medium) and supports the potential applications in clinics.

**Table 1.**
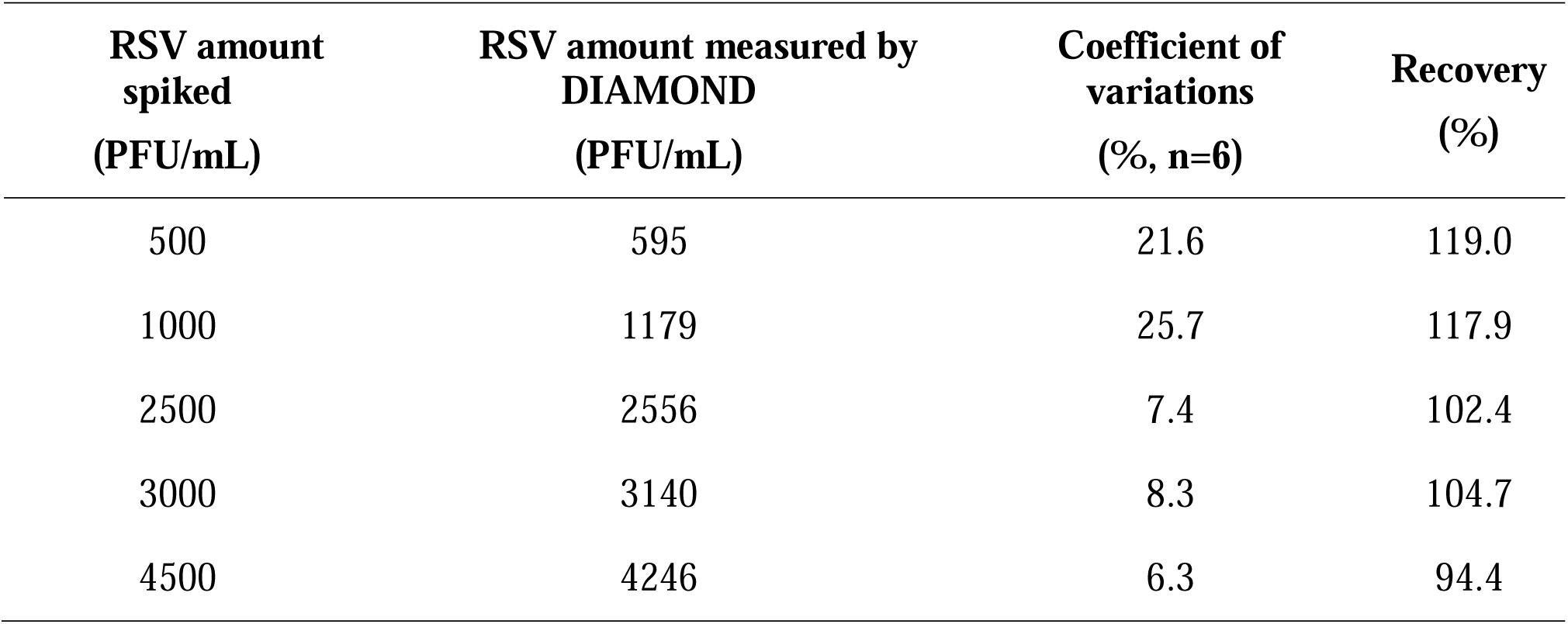
Analytical performance of DIAMOND in detecting RSV-spiked transporting medium.

| **RSV amount**<br>**spiked**<br>**(PFU/mL)** | **RSV amount measured by**<br>**DIAMOND**<br>**(PFU/mL)** | **Coefficient of**<br>**variations**<br>**(%, n=6)** | **Recovery**<br>**(%)** |
| --- | --- | --- | --- |
| 500 | 595 | 21.6 | 119.0 |
| 1000 | 1179 | 25.7 | 117.9 |
| 2500 | 2556 | 7.4 | 102.4 |
| 3000 | 3140 | 8.3 | 104.7 |
| 4500 | 4246 | 6.3 | 94.4 |

## Discussion

Since the PNB generation is dependent on the laser fluence, we can perform the DIAMOND test in two modes by modulating the laser fluence above and below PNB generation threshold, referred to as above- and below-threshold modes. Specifically, the threshold of laser fluence (*E*) was defined as the 50% probability of PNB generation (*P*_PNB_) as determined from the logistic regression (**Fig. S10**) *(20)*. In principle, the below-threshold mode uses low laser fluence at *P*_PNB_=0% (red arrow), resulting in the generation of PNB signals from only the larger NPs or clusters instead of the small NPs or monomers. Alternatively, the above-threshold mode utilizes high laser fluence at *P*_PNB_=100% (blue arrow) to activate all NPs for PNB generation. Despite the background-free signal counting, the below-threshold mode requires an additional step for the laser fluence calibration in order to avoid the incidents where the laser fluence is too low to generate PNB signals from the larger NPs or clusters. In the above-threshold mode, high laser fluence leads to consistent PNB generation that can be visually determined without calibration. However, additional tests for the negative control sample as a reference are necessary to determine the threshold. This can be addressed by splitting the laser beams for simultaneous detection of positive and negative controls, and thus providing automated background-subtracted detection and internal control for validation.

In the digital assays, increasing the total counting number benefits the sensitivity enhancement and reduces the measurement error *(6)*. In the DIAMOND platform, we can increase the data collection time, but this is a tedious way to acquire more data points (**Fig. S11** and **Table S3**). Alternatively, we can use pulse laser with higher repetition rate. Considering the PNB’s lifetime is at nanosecond range, we can use a pulsed laser with repetition rate up to 1 MHz (pulse interval = 1 microsecond). Also, we relied on the repetition rate and flow speed to generate independent PNB events. Taking all parameters (e.g., flow speed, laser diameter, repetition rate, and capillary size) into consideration, we calculated the sampling efficiency of current system is 20-40% along and 5-10% orthogonal to the flow direction, respectively (**Fig. S12**). Such low sampling efficiencies greatly limit the number of detected events and lead to Poisson noise and measurement error. Considering a repetition of 10-100 kHz, a micro-channel with a comparable size to the beam diameter and a faster flow speed, this would allow collecting million nanobubble signals within minutes. We envision that such optimizations may allow further improvements in the performance of DIAMOND in terms of the detection sensitivity and measurement uncertainty.

In summary, we have developed DIAMOND as a novel method for single-particle digital counting in a AuNP homogeneous assay. DIAMOND allows single NPs detection and identification of heterogeneity from a uniform population. Such capability makes DIAMOND a compartment-free digital counting platform and allows its implementation on homogeneous assays. Importantly, DIAMOND achieves enhanced detection sensitivity and completes tests within minutes without additional steps, making it a highly promising platform for rapid and ultrasensitive diagnostics.

## Data Availability

All data generated or analyzed during this study are included in the manuscript (and its supplementary information files).

## Acknowledgments

This research is partially supported by National Institute of Health (NIH) grants R21AI140462 and R01AI151374, and U.S. Department of Defense (DOD) grant PR192581;

## Author contributions

Y.L. and H.Y. carried out the experiment and performed the data analysis. Y.L. H.Y. and H.H. prepared the virus samples. Y.L. P.K. and C.X. developed the MATLAB script. Y.L. H.Y. and Z.Q. wrote the manuscript. Z.Q. and J.S.K. conceived the original idea and supervise the project. H.Y. helped supervise the project. All authors revised the manuscript and have given approval to the final version of the manuscript;

## Supplementary Materials

Materials and Methods

Figures S1-S14

Tables S1-S4

References*(26, 29-32)*

