## Supplemental for "Single-Particle Counting Based on Digital Plasmonic Nanobubble Detection for Rapid and Ultrasensitive Diagnostics"

^#^Co-first authors

**This PDF file includes:**

Materials and Methods

Figs. S1 to S14

Tables S1 to S4

References (*26*, *29-32*)

Materials and Methods

Materials

Tetrachloroauric(III) trihydrate (HAuCl_4_·3H_2_O, 16961-25-4, 99.9%), sodium citrate tribasic dihydrate (Na_3_CA·2H_2_O, 6132-04-3, ≥99%), and hydroquinone (123-31-9, ≥99%), sodium chloride (NaCl, 7647-14-5, ≥99.5%) were purchased from Sigma-aldrich. 3,3'-dithiobis (sulfosuccinimidyl propionate) (DTSSP, 21578, 50 mg) and borate buffer (1M, 28341) were purchased from Thermo Scientific. Sucrose (57-50-1), Magnesium sulfate hydrate (MgSO4, 22189-08-8), HEPES (7365-45-9), Dulbecco's modification of eagle's medium (EMEM), fetal bovine serum (FBS), serum-free medium (SFM), hydrogen chloride (HCl, 7647-01-0), ethanol (64-17-5, 190 proof), and Amicon™ ultra centrifugal filter units (UFC510024) were purchased from Fisher Scientific. Aminated SiO_2_ beads in ethanol (SIAN500-25M) were purchased from Nanocomposix. Anti-RSV fusion protein antibody, palivizumab/Synagis was obtained from MedImmune, Gaithersburg, MD, CAT # NDC60574). HEp-2 cells were purchased from American Type Culture Collection (ATCC), CAT # CCL-23. All aqueous solutions were prepared using deionized (DI) water (dH_2_O) with a resistivity of 18.0 MΩ·cm.

Methods

Nanoparticle synthesis and characterization

AuNP seeds were firstly synthesized using classical Plech Turkevich’ method with slight modifications (*29*). Briefly, 1 mL of HAuCl_4_ aqueous solution (25 mM) was added to a clean 250 mL Erlenmeyer flask with 98 mL of pure water and allowed to boil on a hot plate with magnetic stirring. Subsequently, 1 mL of sodium citrate (112.2 mM) aqueous solution was quickly injected into the boiling solution with a pipette. The solution was kept stirring and boiling for 10 min until its color turned red. After cooling the solution to room temperature, dH_2_O was added to bring the volume to 100 mL. The products were stored in the dark at room temperature for future use.

Large size of AuNPs were synthesized according to a seed-mediated growth method with slight modifications, where above-mentioned AuNPs were employed as the seeds. In brief, the appropriate amount of pure water, HAuCl_4_ precursor, sodium citrate, and AuNP seeds were added orderly to a clean 250 mL flask under vigorous magnetic stirring at room temperature according to **Table S4**. Then the reducing aqueous solution containing a certain amount of hydroquinone was injected rapidly to the flask. The solution immediately switched color to purple and then to red in a few minutes. The reaction was left overnight at room temperature. Final products were stored in the dark at room temperature for future use. Note that all particles concentration in this study were determined based on the size-dependent empirical formula with a combination of UV-vis measurement and transmission electron microscope (TEM) (*26*).

Biosafety statements

The research project was approved and performed strictly in adherence to CDC/NIH guidelines and the experimental protocols were approved by the University of Texas at Dallas Institutional Biosafety & Chemical Safety Committee and the University of Texas Southwestern Medical Center Biosafety Committee. Human Metapneumovirus (hMPV, CAT# 0810163CF) and Parainfluenza virus type 1 (PIV, CAT# 0810014CF) were purchased from ZeptoMetrix, and H1N1 influenza virus type A (IVA, CAT# IHA-003) was purchased from ProSpec-Tany TechnoGene Ltd. Human respiratory syncytial virus (RSV) A2 strain (CAT # VR-1540) was purchased from ATCC.

Conjugation AuNPs with Synagis as detection probes for RSV

Conjugation of AuNPs with Synagis was adapted from a previous report (*30*). In brief, 5 mM DTSSP as a crosslinker was reacted with primary amines of antibody at a molar ratio of 125:1 in 2 mM borate buffer first, then followed by an overnight membrane dialysis process to eliminate free DTSSP before concentrated using 100 kDa Amicon centrifugal filter. The resulted product (DTSSP-Syn) was then mixed with 15 nm AuNPs suspension in 2 mM borate buffer at a molar ratio of 500: 1 and incubated overnight at 4 °C. Afterwards, the products (15 nm AuNP probes) were washed three times with 2 mM borate buffer and re-dispersed in 2 mM borate buffer and kept at 4 ^o^C for storage before further testing.

Large scale propagation of RSV A2 in HEp-2 cells

RSV A2 strain was propagated in HEp-2 cells. HEp-2 cells were cultivated in EMEM/5% FBS and infected at a multiplicity of infection (MOI) of 0.01 to minimize the production of defective interfering (DI) particles. Viral working stocks were prepared using 30%-60% (w/v) non-continuous sucrose density centrifugation (Beckman Coulter SW-28 rotor, 28000 rpm, 4°C for 90 minutes). The virus containing band at the 30%-60% interface was collected (**Fig. S13**), distributed into aliquots, and stored at -80 ^o^C *(31)*. Viral titers were determined by endpoint dilution assay (**Fig. S14**) as previously described *(32)*.

Detection of RSV using the AuNP-based probes

RSV and other closely related respiratory viruses such as hMPV, PIV, and IVA were then incubated with AuNP probes for at least 30 min at room temperature in the presence of 5% sucrose and cell debris in 1×MHS buffer (1 M MgSO_4_, 0.5 M HEPES, pH=7.7, 1 M NaCl).

PNB generation and detection

The PNB generation was conducted using ultrafast pump laser pulses (28 ps, 532 nm, PL2230, Ekspla). The AuNPs suspension was pushed through a 200 μm micro-capillary (VitroTube, 8320) using a syringe pump (New Era Pump Systems Inc.) and irradiated by pump laser. The laser fluence was varied by rotating the beam attenuator (ThorLABs, VBA05-532) and was measured using a laser power meter (FieldMaxII-TOP, Coherent). PNB signals were monitored by a continuous laser as probe beam (Red HeNe laser, 633 nm, R-30989, Newport) and its intensity was recorded with a photodetector (FPD510-FV, Thorlabs) and an oscilloscope (LeCroy WaveRunner204Xi-A).

For the 75 nm AuNPs detection, serial suspensions with concentrations equaling λ=80, 40, 27, 8, 4, 2.7, 0.8, 0.4, 0.27, 0.08, 0.04 were prepared and tested at laser fluence of 10,000 mJ/cm^2^. All the measurements were recorded one minute per sample (3,000 pulses at 50 Hz).

For the different size detection, serial suspensions of AuNPs with 15 nm, 35 nm, 50 nm, and 75 nm in diameter and 1.5×10^10^ mL^-1^ (λ=240) in concentration were prepared and tested at laser fluence of 2,000 mJ/cm^2^.

For the mixture sample test, the particle concentrations of AuNP suspensions were set the same as 1.5×10^10^ mL^-1^ (λ=240) and 3.125×10^7^ mL^-1^ (λ=0.5) for 15 and 75 nm AuNPs, respectively. The PNB detection was performed at laser fluence of 7,500 mJ/cm^2^.

For the SiO_2_ beads detection, the as-purchased SiO_2_ beads in ethanol was first centrifuged and washed with dH_2_O twice and re-dispersed in 2 mM citrate-HCl buffer (pH=3) with particle concentration of 1×10^10^ mL^-1^. The SiO_2_ beads were sonicated and vortexed before mixing with 15 nm citrate-AuNPs suspension. The mixture was incubated for 30 minutes at room temperature before further tests. The PNB detection was performed at laser fluence of 3,000 mJ/cm^2^.

For the RSV detection, the completed assay solutions were directed to the DIAMOND test at laser fluence of 3,000 mJ/cm^2^.

MATLAB functions

The raw data of PNB signals were processed by a customized MATLAB script. The major functions of the script include pre-filtration, PNB signal recognition by signal-to-noisy-ratio (SNR), parameters extraction, threshold calculation, and frequency counting for “on” signals.

Characterization *techniques and data analyses*

The absorbance of samples in microtiter plates was read using microplate reader (Synergy 2, BioTek). The DLS was measured using Malvern ZetaSizer Nano ZS. Extinction spectrum were obtained with a spectrophotometer (DU800, Beckman Coulter). The TEM images were taken using a JEOL JEM-2010 microscope operated at 120 kV. The pH values of buffer solutions were measured using a pH Meter (Accumet AP71). All data were collected in no less than triplicate and reported as mean and standard deviation for statitical analysis. Two-sample t-test assuming equal variance in Origin software was conducted to determine *p* values and statistical significance.

Poisson statistics

The theoretical prediction by Poisson statistics in our study was calculated by following equation:

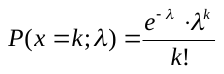

where *k* and *λ* are the actual and average number of AuNPs in compartments, respectively, and *e* is Euler’s number. The data in **Fig. S4** was generated using Microsoft Excel Worksheet (Data-Data Analysis-Random Number Generation-Poisson).

**Table S1. Comparison of various digital assays.**

|  | **Sensing format** | **Sample partition** | **Label** | **Signal amplification** | **Readout** | **Reference** |
| --- | --- | --- | --- | --- | --- | --- |
| **Erenna** | ^a^Bead-based ELISA | Laser confocal/Capillary electrophoresis | Fluorescence | No | Counting | 5 |
| **dELISA** | Bead-based ELISA | Microwells array | Enzyme | Yes | Imaging | 3, 7 |
| **dPCR** | ^b^PCR | Droplets | Nucleic acids | Yes | Counting | 4 |
| **Microscopy** | Bead-based ELISA + PCR | Individual isolates | Magnetic beads | Yes | Imaging | 6 |
|  | ^c^Homogeneous assay | Microwells array | Magnetic beads | No | Particle motion | 17, 18 |
|  |  | Individual isolates |  |  |  | 19 |
|  | Homogeneous assay | Individual isolates | Plasmonic NPs or Quantum dots | No | Imaging | 14, 16 |
|  | ^d^Heterogeneous sandwich assay |  |  |  |  | 11-13, 15 |
| **DIAMOND** | Homogeneous assay | Laser confocal/ Capillary | Plasmonic NPs | No | Counting | This work |

^a^: Bead-based ELISA means performing the enzyme-linked immunosorbent assay (ELISA) on individual beads. Typical procedures include conjugation of primary capture antibody on beads, linkage of enzyme on a secondary antibody, and incubation of antigen with both antibodies. Multiple cycles of washing steps are required during each step to remove the impurities. Amplification is also need.

^b^: PCR means polymerase chain reaction, where the target needs to be amplified prior to the measurement. Primers and temperature control are needed for specific amplification. Similar work includes digital loop-mediated isothermal amplification and digital clustered regularly interspaced short palindromic repeats-based detection.

^c^: Homogeneous assay means using one type of labels to conjugate single target, followed by measuring without additional steps.

^d^: Heterogeneous sandwich assay sensing format means using two types of labels to conjugate single target with one label pre-immobilized on a surface, followed by washing steps to remove the other label in free and then measuring the signals.

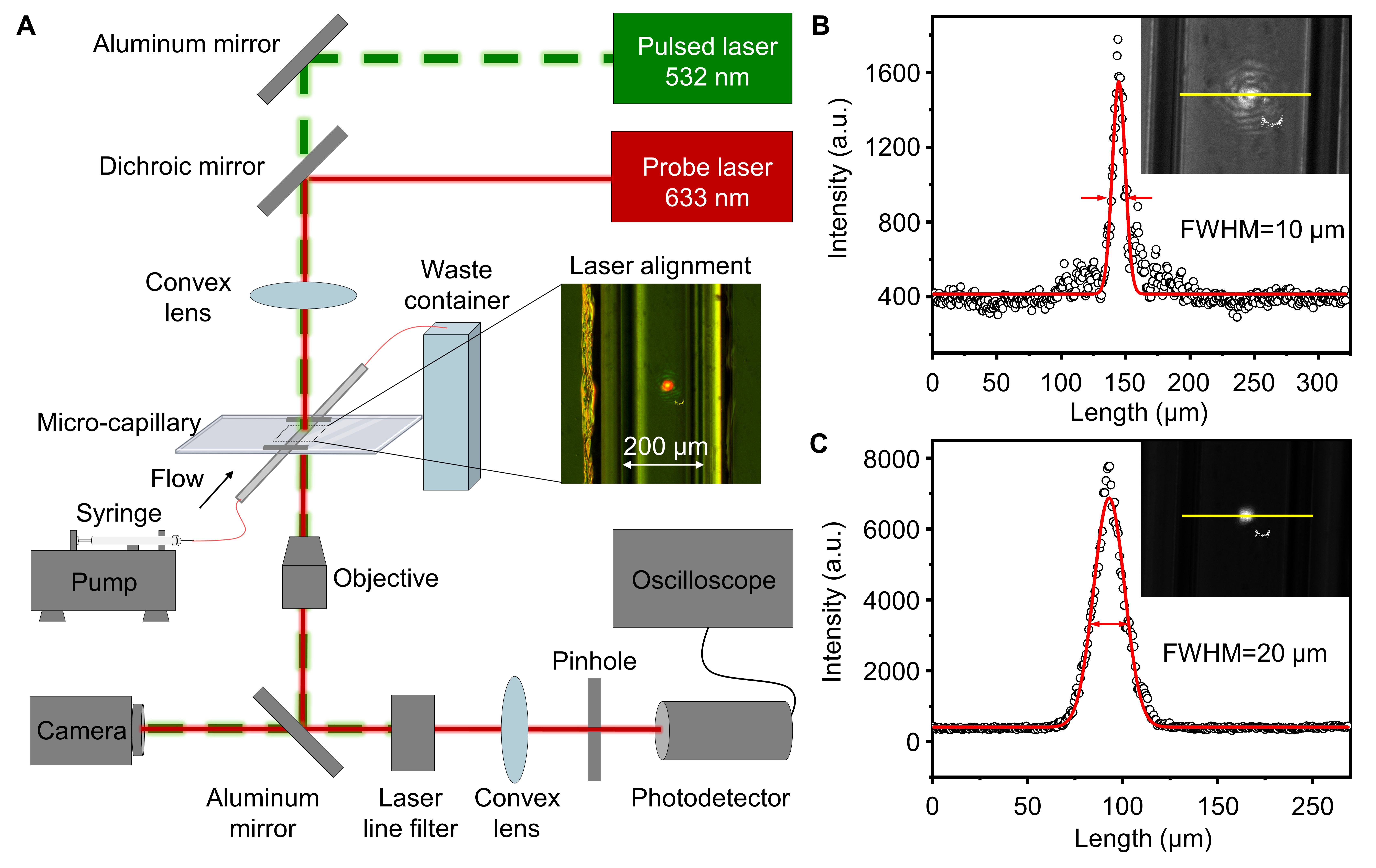

Fig. S1. DIAMOND apparatus and laser beam characterization. (A) Schematics illustration of the experimental setup. Energy profiles of (B) pump and (C) probe lasers. The scatters were normal distribution fitted, where the full width at half maximum (FWHM) was used as beam diameters. The white pattern in insets is a burned damage to the camera.

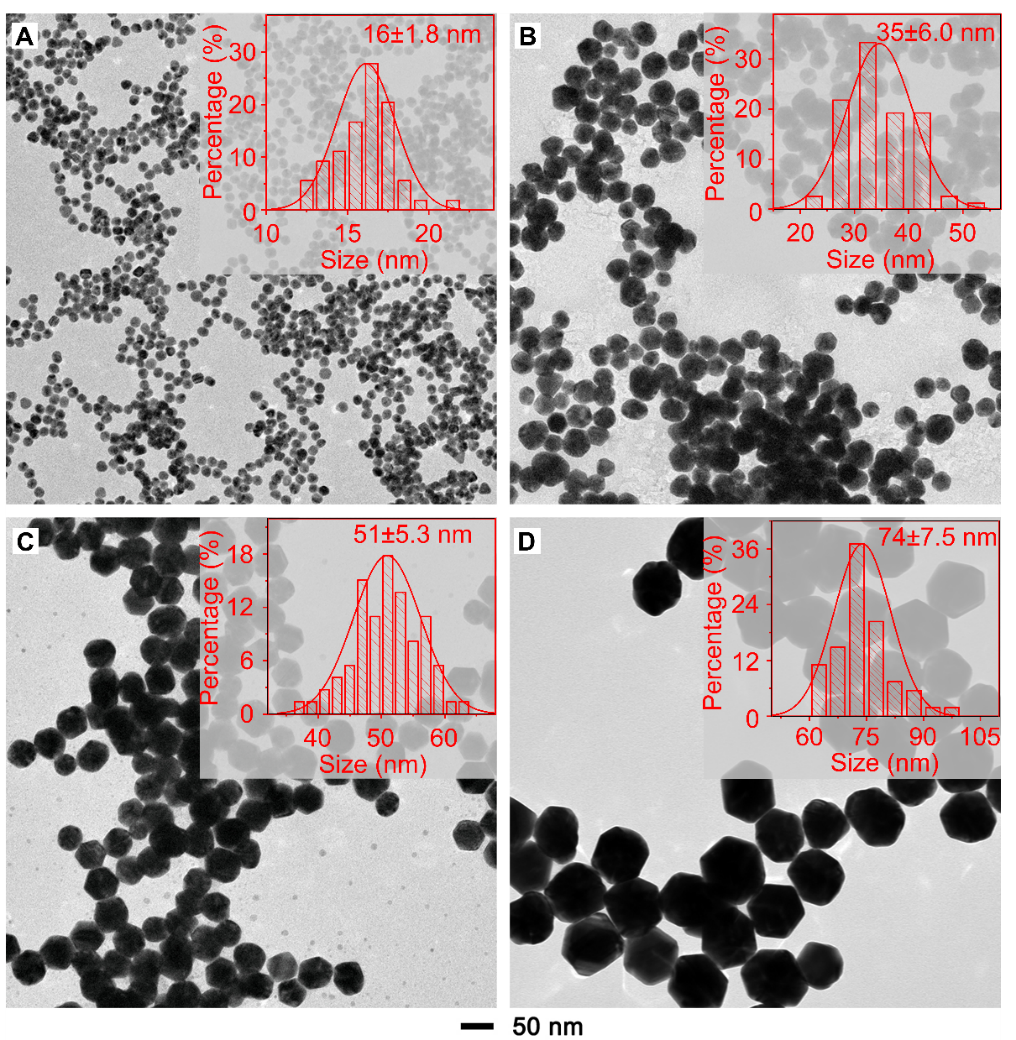

Fig. S2. Morphology and size distribution histograms of different AuNPs by TEM. The results were counted by randomly measuring 200 particles in TEM images. For simplicity, we referred the AuNPs in (A-D) as 15 nm, 35 nm, 50 nm, and 75 nm, respectively, and used them through the whole manuscript.

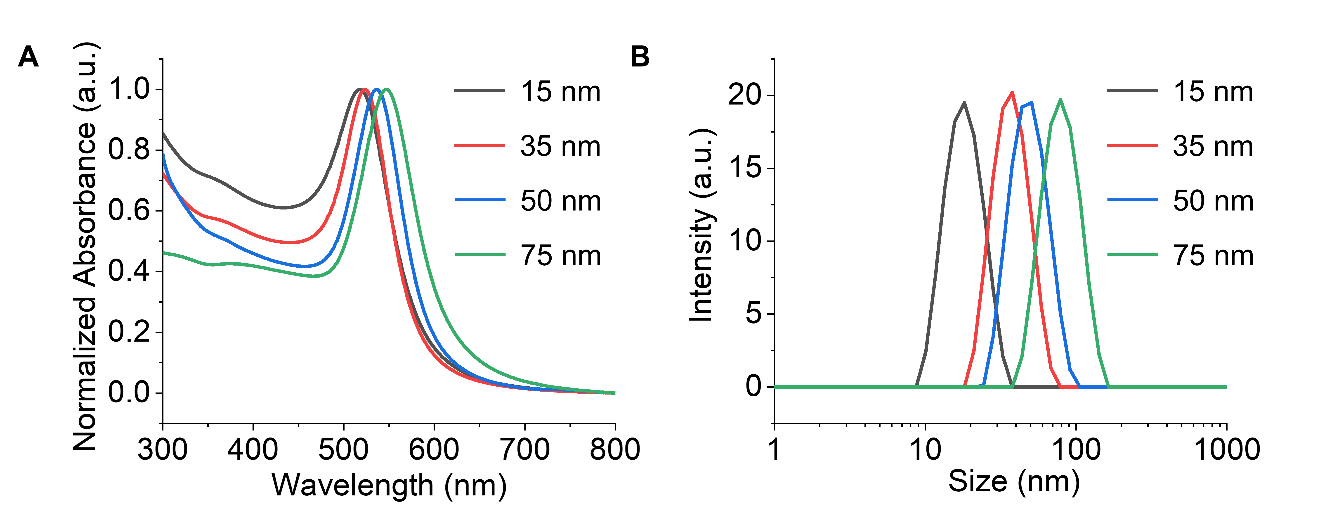

Fig. S3. Optical characterizations for AuNPs of different sizes. (A) UV-Vis measurements. (B) DLS analysis.

Table S2. Summary of the characterizations of AuNPs used in the study.

| AuNP size (nm) | TEM | | UV-Vis spectra | DLS analysis | |
| --- | --- | --- | --- | --- | --- |
|  | ^a^Diameter (nm) | Standard deviation (nm) | Peak location  (nm) | Diameter (nm) | Polydispersity index (PDI) |
| 15 | 16 | 1.8 | 518 | 17.3 | 0.062 |
| 35 | 35 | 6.0 | 524 | 38.1 | 0.062 |
| 50 | 51 | 5.3 | 537 | 55.2 | 0.067 |
| 75 | 74 | 7.5 | 547 | 76.2 | 0.044 |

^a^: Measured by counting 200 nanoparticles randomly.

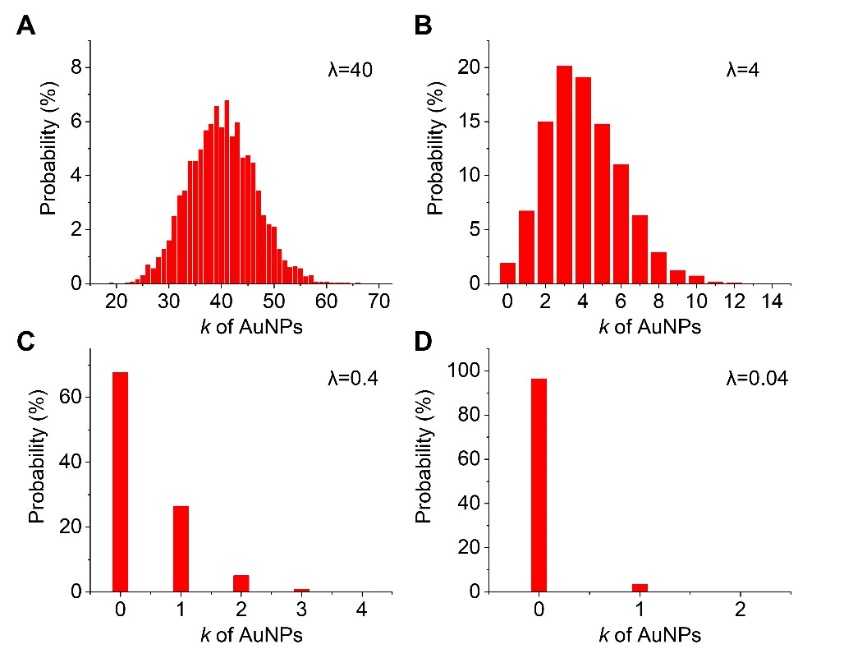

Fig. S4. Probability distribution histograms of the AuNP number (*k*) in each virtual compartment for a given λ, based on Poisson statistics.

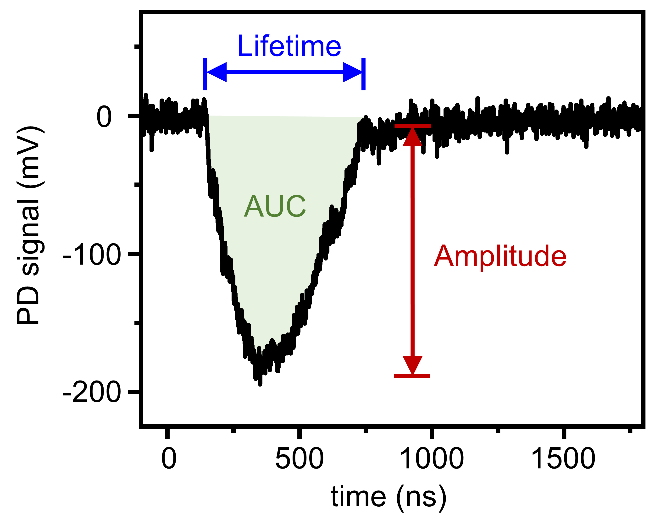

Fig. S5. Schematic illustration of the indexing parameters for a photodetector (PD) signal. It includes the amplitude (peak intensity), lifetime (peak width), and area-under-curve (AUC, peak area). In the present work, only values of amplitude and AUC were used as indexes.

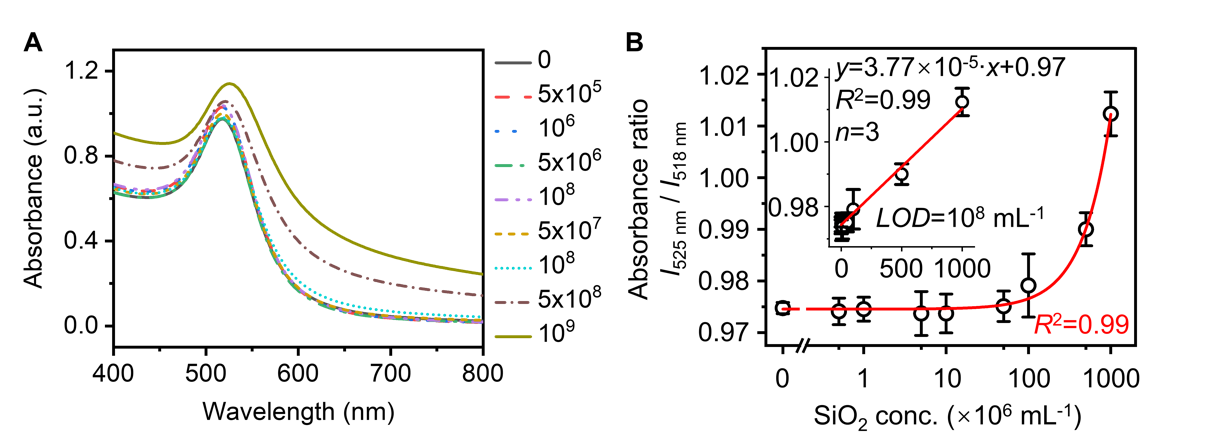

Fig. S6. Colorimetric detection of homogeneous assay using AuNPs as probes and SiO_2_ beads as targets. (A) Absorbance spectra of assay solutions with different SiO_2_ beads concentrations (particles/mL). (B) Colorimetric analysis of the detection result. Inset shows the linear range of the colorimetric detection. Error bars indicate the standard deviations of three independent measurements, and the *LOD* was calculated as 3 standard deviations of the control divided by the slope of regression line.

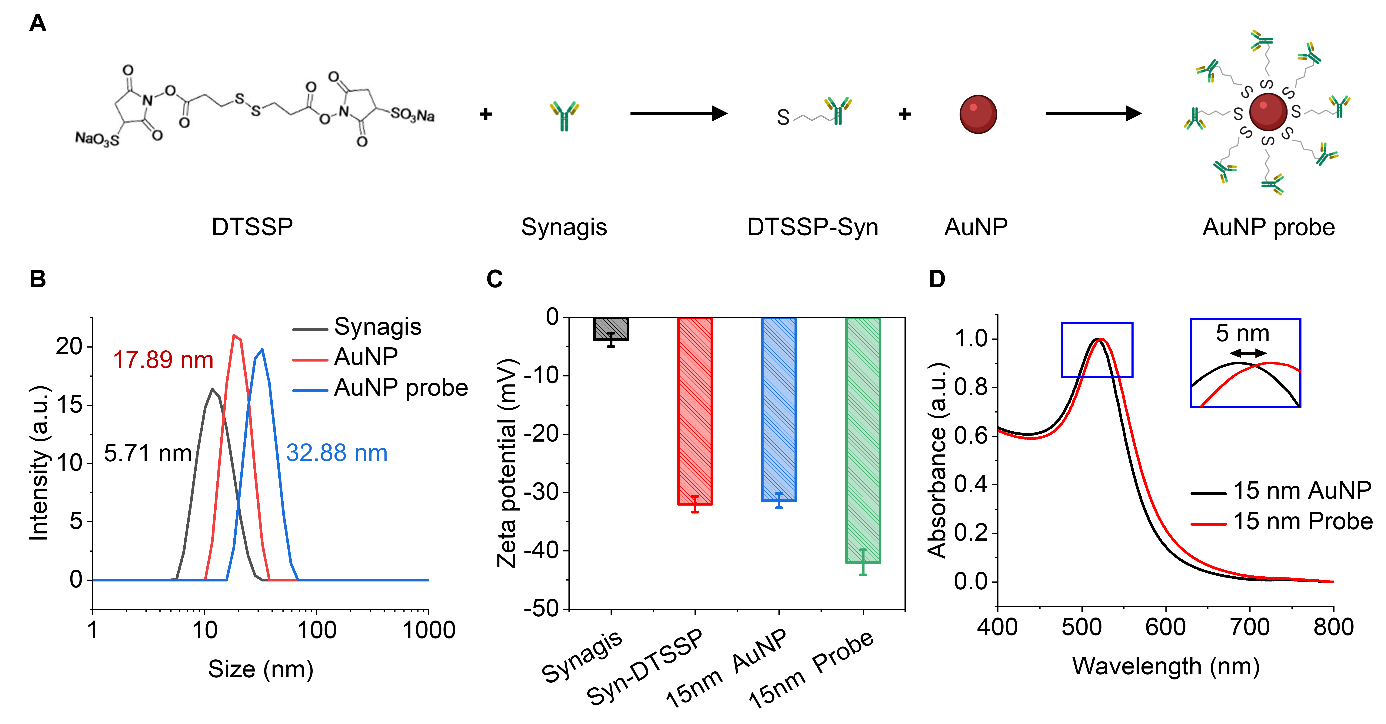

Fig. S7. Preparation and characterization of AuNP-based probes for the RSV detection. (A) Schematic illustrates the preparation of 15 nm AuNP probes. (B) DLS, (C) Zeta potential, and (D) UV-Vis measurement for the AuNP probes characterization before and after conjugating with antibody-linked DTSSP.

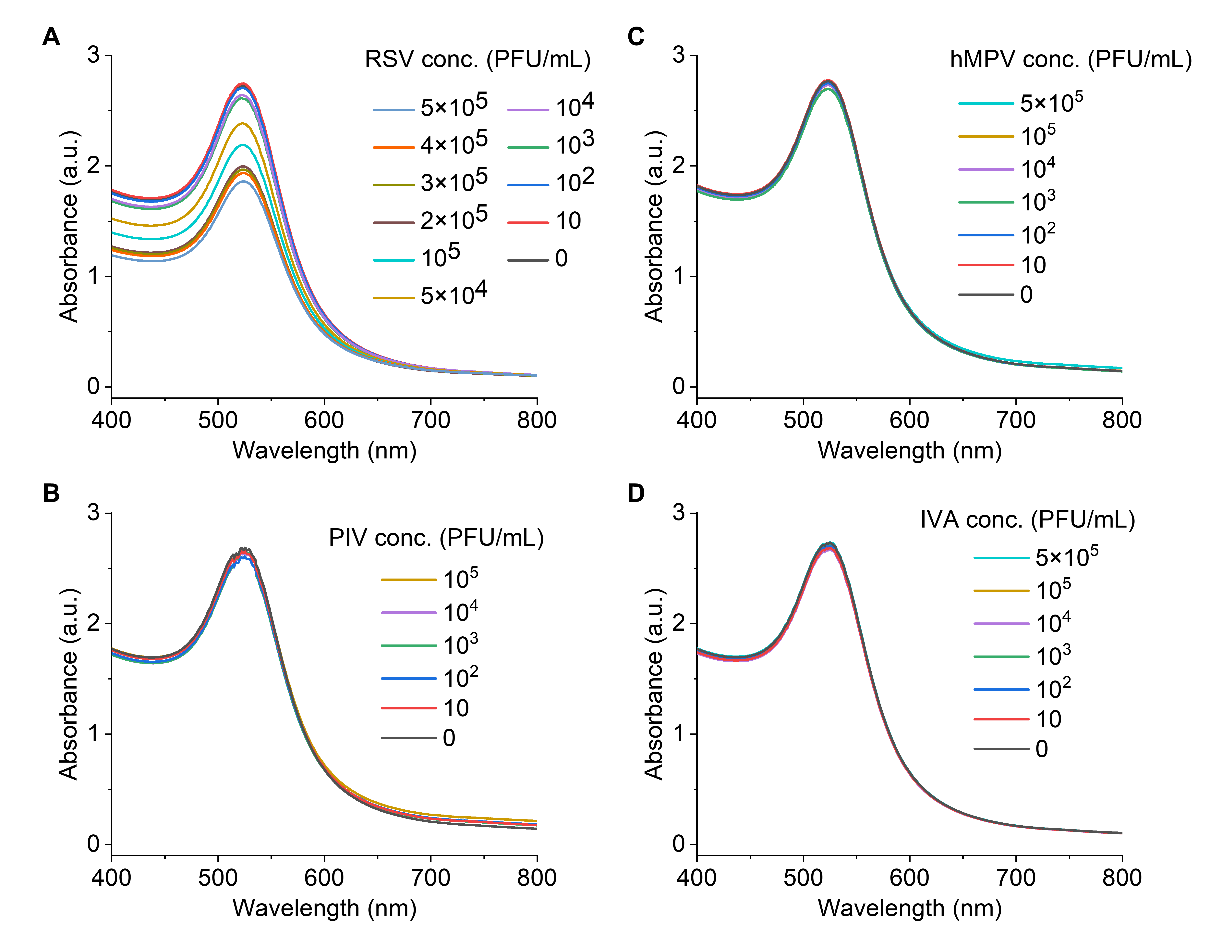

Fig. S8. Absorbance spectra monitoring the homogeneous immunoassays of different respiratory viruses with varied titers. (A) RSV=respiratory syncytial virus. (B) PIV=Parainfluenza viruses. (C) IVA=Influenza A. (D) hMPV=Human metapneumovirus.

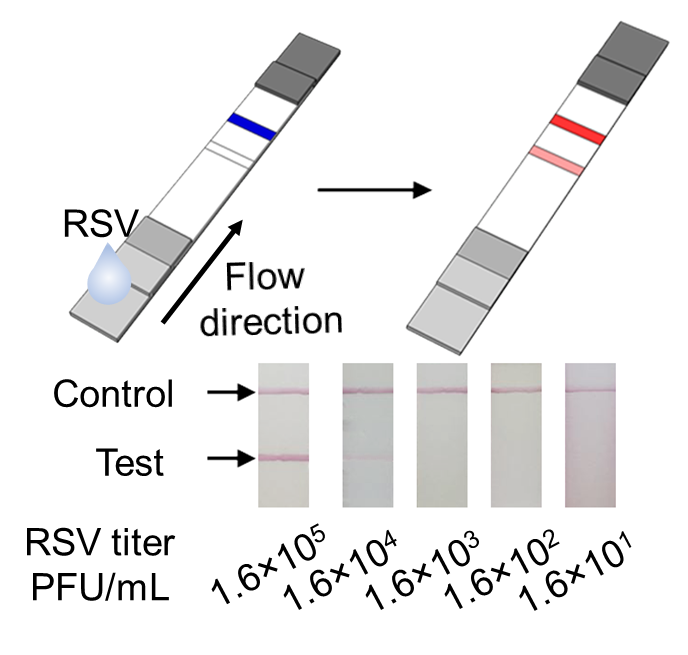

Fig. S9. RSV detection using a commercial lateral flow assay kit (BinaxNOW, Abbott). Schematic illustrates the assay operation and the digital photographs show the corresponding detection result with different RSV titers. The detection limit was estimated to be 1.6×10^4^ PFU/mL.

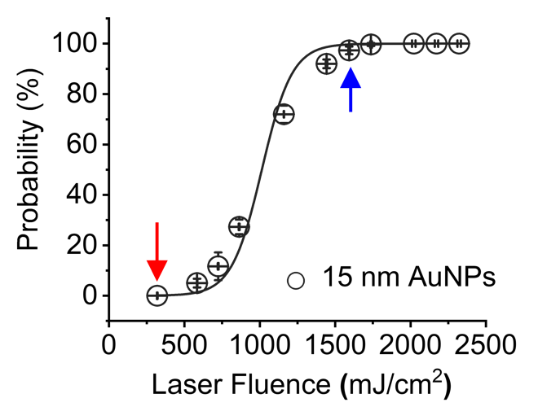

Fig. S10. Example of the PNB generation probability curve of 15 nm AuNPs used to determine the laser fluence threshold. Red arrow marks the laser fluence for probability=0, while the blue arrow marks the laser fluence for probability =100%.

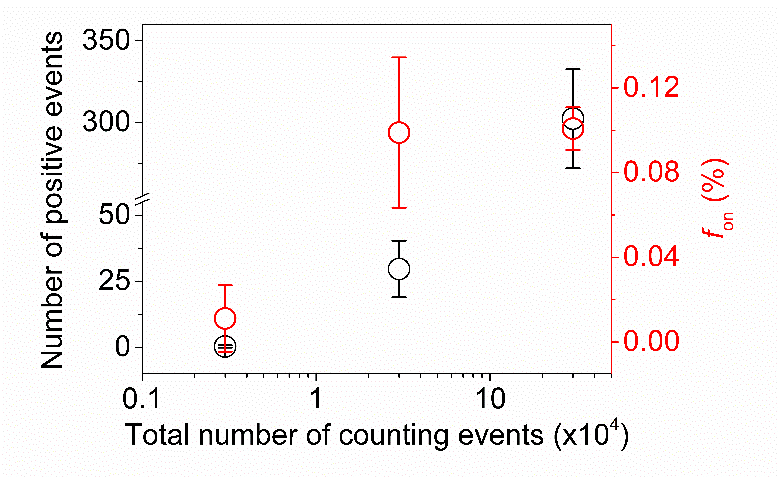

Fig. S11. The detection performance for the detection of 75 nm AuNPs (λ=0.0004) by DIAMOND with increasing counting number.

Table S3. A prediction on the sensitivity enhancement by means of increasing the counting number for DIAMOND.

| ^a^Expected particle number (λ) | Poisson prediction (%) | Average positive counts of 1 thousand counts | Average positive counts of 1 million counts |
| --- | --- | --- | --- |
| 0.1 | 9.5 | 95 | 95,000 |
| 0.01 | 1 | 10^*^ | 10,000 |
| 0.001 | 0.1 | 1 | 1,0002 |
| 0.0001 | 0.01 | 0.1 | 100 |
| 0.00001 | 0.001 | 0.01 | 10^*^ |
| 0.000001 | 0.0001 | 0.001 | 1 |

^a^: λ is proportional to the target concentration for a given detection volume

^*^: estimated *LOD*

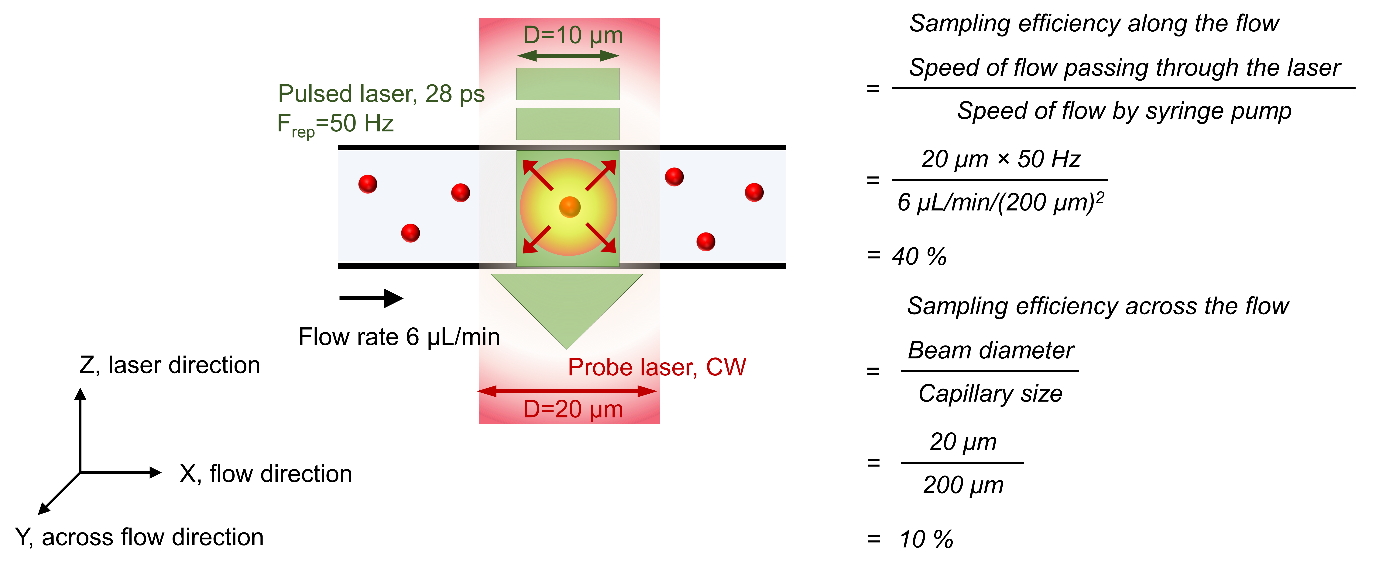

Fig. S12. Schematic illustrates the calculation of sampling efficiency for the probe beam based on setup configuration. Note the corresponding sampling efficiency of pump beam can also be calculated as 20% and 5%, respectively.

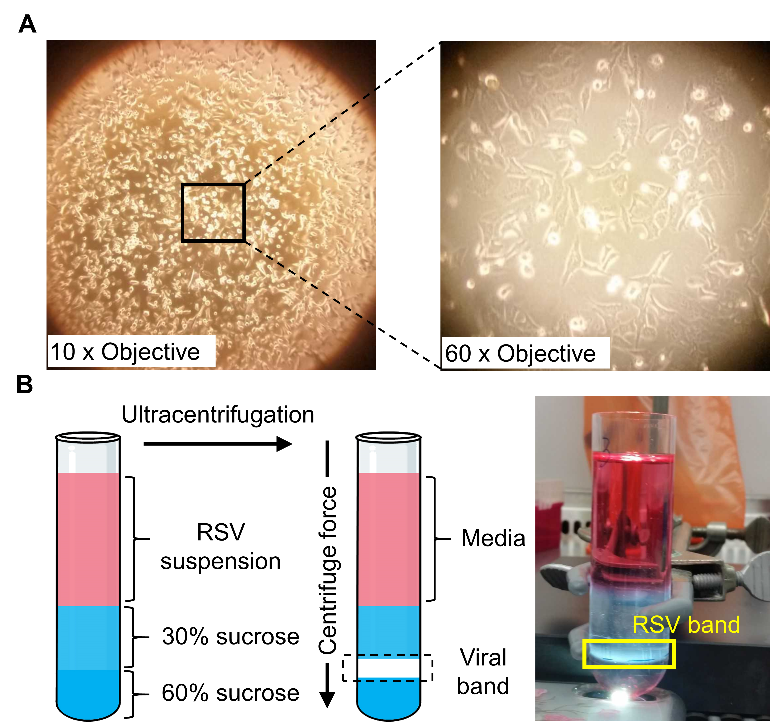

Fig. S13. Large scale preparation of RSV A2. (A) Photographs of clinical RSV A2 strain infected HEp-2 cells at day 4. (B) Sucrose density gradient for RSV purification.

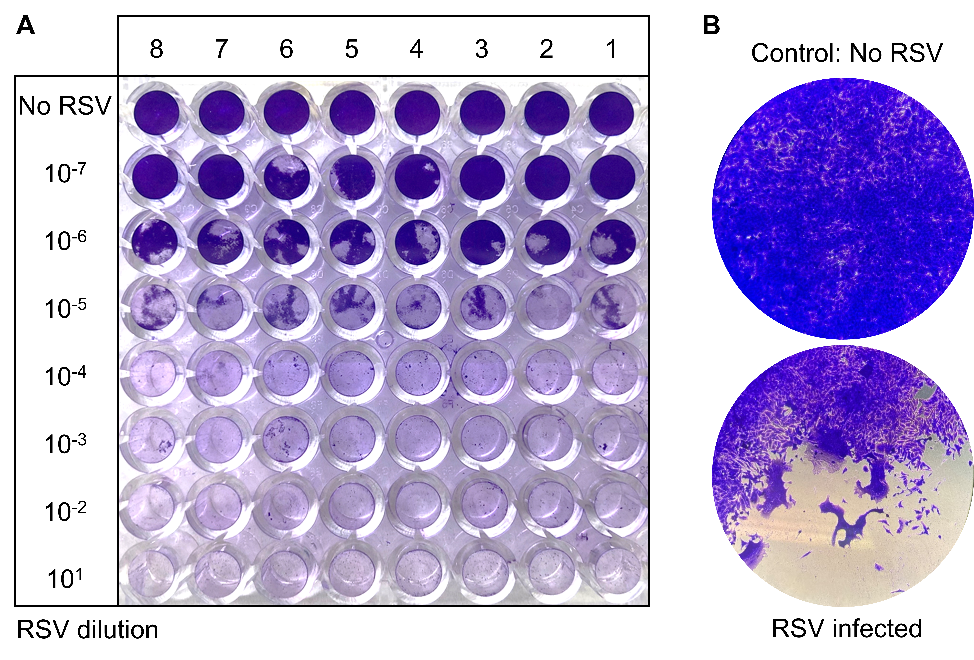

Fig. S14. Endpoint dilution assay for RSV quantification. (A) 96 well plate layout for serial dilutions of RSV stock inoculated onto HEp-2 cells. (B) Example of control well and RSV infected well after staining with crystal violet dye.

Table S4. One-pot recipe for the synthesis of AuNPs based on seed-growth method.

| Reagent  Size (nm) | H_2_O  *mL* | HAuCl_4_  *25 mM, mL* | Sodium Citrate  *15 mM, mL* | 15 nm Au Seeds  *2.23 nM, mL* | Hydroquinone  *25 mM, mL* |
| --- | --- | --- | --- | --- | --- |
| 35 | 84.84 | 0.875 | 0.875 | 12.533 | 0.875 |
| 50 | 93.39 | 0.963 | 0.963 | 3.714 | 0.963 |
| 70 | 96.67 | 0.997 | 0.997 | 0.338 | 0.997 |
